# A Pilot Comparative Study of Dental Students’ Ability to Detect Enamel-only Proximal Caries in Bitewing Radiographs With and Without the use of AssistDent^®^ Deep Learning Software

**DOI:** 10.1101/2020.06.15.20131730

**Authors:** Hugh Devlin, Martin Ashley, Tomos Williams, Brian Purvis, Reza Roudsari

## Abstract

Enamel-only proximal caries, if detected, can be reversed by non-invasive treatments. Dental bitewing radiograph analysis is central to diagnosis and treatment planning and when used to detect enamel-only proximal caries it is an important tool in minimum intervention and preventive dentistry. However, the subtle patterns of enamel-only proximal caries visible in a bitewing radiographs are difficult to detect and often missed by dental practitioners. This pilot study measures the ability of a cohort of third-year dental students to detect enamel-only proximal caries in bitewing radiographs with and without the use of a deep learning assistive software AssistDent^®^. We demonstrate an increased ability in the detection of enamel-only proximal caries by the students using AssistDent, showing a mean sensitivity level of 0.80 (95%CI ± 0.04), increased from 0.50 (95%CI ± 0.13) p<0.01 shown by students not using AssistDent. This improvement in ability was achieved without an increase in false positives. Mean false positives per bitewing radiograph recorded by students when using AssistDent was 2.64 (95%CI ± 0.57), and by students without using AssistDent was 2.46 (95%CI ± 1.51). Based on these results we conclude that the AI-based software AssistDent significantly improves third-year dental students’ ability to detect enamel-only proximal caries and could be considered as a tool to support minimum intervention and preventive dentistry in teaching hospitals and general practice. We also discuss how the experience of conducting this pilot study can be used to inform the design and methodology of a follow-on study of AssistDent in dental practice use.

## Introduction

Two recent meta-analysis studies have found that dentists using visual and clinical examination and radiographs have a poor sensitivity in detecting proximal caries but have good specificity [1] [2]. Mean sensitivity varies across different studies, but some studies report that less than 50% of the enamel and dentine caries is detected with combined visual examination and bitewing radiography [3]. Other studies have shown that this sensitivity is reduced further when subjects are asked to diagnose enamel caries alone. Subjects with greater experience make more accurate assessments [4], so training is important in preventing misdiagnosis and overtreatment. However, in one study, almost two thirds (61%) of the proximal caries found by the specialists failed to be detected by experienced general dentists [5]. Many other factors influence whether a carious lesion is detected by a dentist such as the viewing conditions, quality of radiographs and the presence of artefacts and the size of the lesion on the radiograph.

Early detection of caries prior to surface enamel cavitation is necessary for effective minimal intervention and preventive dentistry. Once an individual’s caries risk is assessed accurately, their caries risk factors can be analysed, and effective prescription and delivery of preventive strategies initiated. Enamel-only proximal caries remineralizes more readily than larger lesions [6]. If the intervention fails to prevent caries, then the clinician will need to detect this at the earliest opportunity.

Dental undergraduates need to have cariology detection and management systems integrated into their clinical patient assessment and treatment. Tikhonova [7] described a situation in all Canadian dental schools where effective integration of cariology education into clinical training was not being implemented. For example, only one dental school used the concept of caries risk assessment to inform their clinical decisions. One solution could be to provide the opportunity to generate electronic dental records that allow the automatic detection of the position of proximal carious lesions.

This study is primarily concerned with the detection of enamel-only proximal caries, and not proximal dentine caries, occlusal or secondary caries.

## AssistDent^®^ AI-Based Detection of Proximal Caries

AssistDent® is a software product developed by Manchester Imaging Limited that uses machine learning algorithms to prompt the dentist by highlighting regions of interest within a bitewing radiograph which are indicative of enamel-only proximal caries. It is intended to be used to aid clinical decision making at the chair-side and as support when discussing treatment options with patients. The final decision about whether an enamel-only proximal caries is present, or not, rests with the dentist as they have access to all the clinical information.

AssistDent is accessed via a web-page graphical user interface. This provides the means for users to log into the system, upload bitewing radiographs, interact with the images, record their clinical assessment and save their final assessment to a central database. Once logged in, users are invited to load a dental bitewing for clinical analysis by dragging it into the interface. Once uploaded, the image appears on the web page and the graphical user interface tools allow users to interact with the image and the AssistDent prompts as shown in Figure 1.

**Figure 1.**
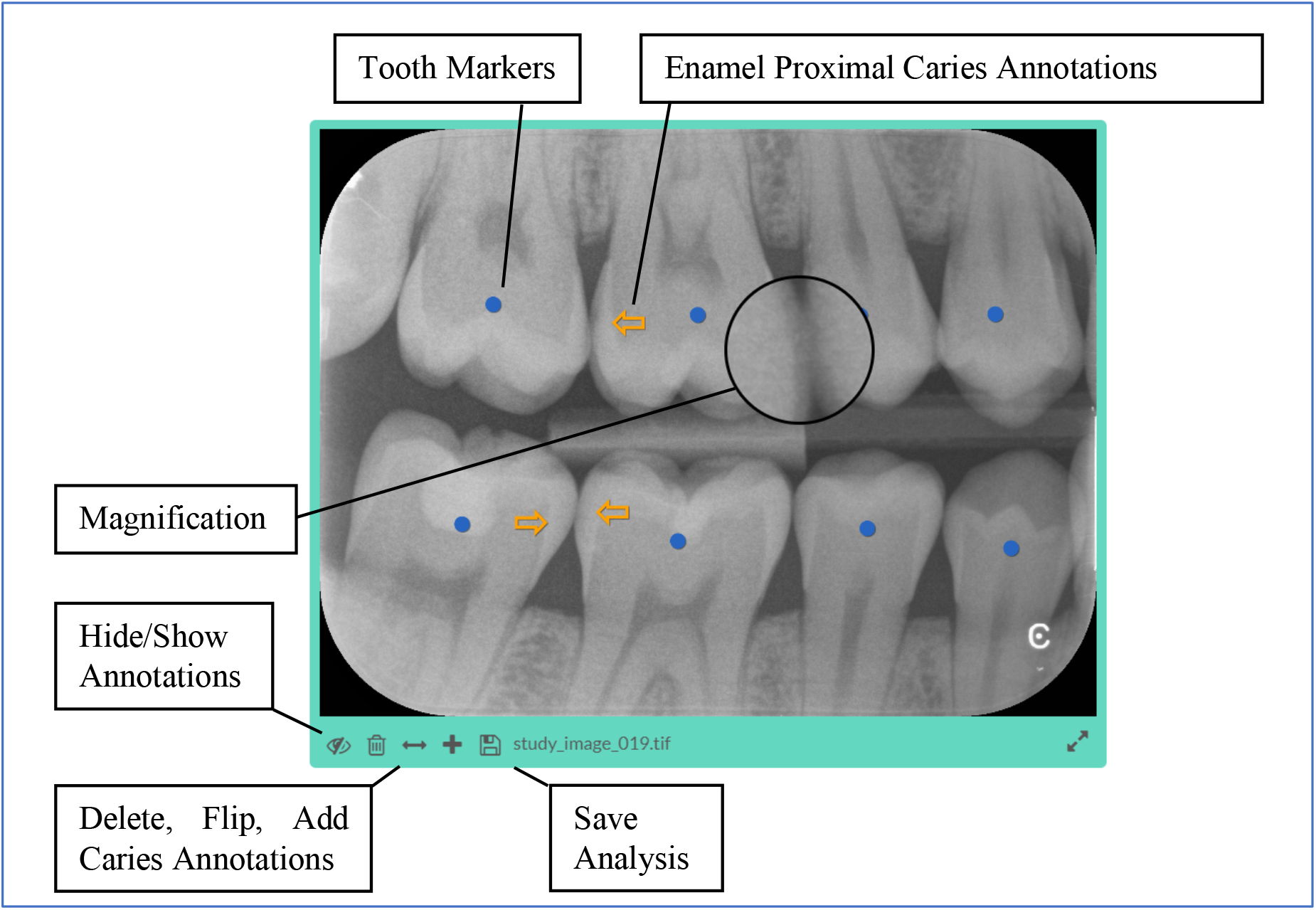
AssistDent Graphical User Interface (GUI). The GUI provides features to assist in the clinical analysis of the bitewing radiograph via a magnifying tool, an ability to display the image using the full screen and to hide any annotations which may obscure pathology. Teeth detected by the AssistDent algorithms are annotated by blue circles in their centres. Regions where AssistDent prompts, or the user believes, that enamel-only proximal caries are present are annotated with orange arrows pointing away from the centre of the tooth towards the tooth edge where the caries is located. Users can edit the caries annotations by adding additional arrows, altering the position and direction of existing arrows and deleting arrows. The user is instructed to save their analysis by selecting the save button which sends the location of all the annotations together with the user and image identification to a central database.

A secondary element of AssistDent is the bitewing analysis. This element constitutes the Artificial Intelligent (AI) element of the software. It consists of a pipeline of machine learning algorithms that detect teeth; find the proximal tooth edges then process these edges to determine regions which are indicative of enamel-only proximal caries. The machine learning algorithms were trained using a set of more than 1,400 bitewing radiographs collected from 1 UK teaching hospital site and 9 UK general dental practices covering a range of intra-oral X-ray units from different manufacturers. Ethical approval, separate to the study, was obtained for a cross-sectional, non-intervention study that involved the collection of anonymized digital bitewing radiographs from digital scanner systems commercially available in the UK (ref 18/NI/0111). Manual annotation of the edges of approximately 3,500 teeth was used to train the tooth detection and tooth edge finding portions of the algorithms.

## Methods

This pilot study was conducted at an early phase in our research process with the purpose of evaluating a novel application of AI-based computer aided diagnosis in dentistry training and inform the design and methodology of a larger scale follow-on study in dental practice [8].

The protocol, participant information sheet and consent forms for the study were approved by the Manchester University Research Ethics Committee (Ref: 2019-8534-12770). The participants were chosen from a cohort of dental students in the third year of their undergraduate course (which in the UK has a duration of five years). Although fairly inexpert at caries detection, the students had commenced clinical caries diagnosis and intracoronal restorative treatment in the undergraduate clinic.

A cohort of 24 students volunteered to participate and were randomly divided into control and experimental arms. One control arm participant failed to analyse any of the images due to technical issues with their home computer. Both arms examined the same images using the same graphical user interface. In the control arm, the caries detection function of AssistDent was disabled in order to measure the enamel-only proximal caries detection ability of participants without the use of assistive software. In the experimental arm, the caries detection function of AssistDent was enabled in order to assist the caries detection ability of participants with assistive software.

A total of 1,446 bitewing radiographs were collected from 10 sites (1 teaching hospital and 9 general dental practices) each with different X-ray acquisition systems for the purposes of algorithm training and evaluation in this study. A validation set of 103 images were selected by random stratified sampling partitioned over the image acquisition sites and excluded from all machine learning model training and evaluation. A further subset of 24 images from the validation set were selected for the study, again stratified over the acquisition sites but with the criterion that there was at least one enamel-only proximal caries in each image. Images from one of the GDP sites were excluded due to their poor quality Insufficient number of images met this criterion for one of the other GDP sites hence two study images from one site had no enamel-only proximal caries. The images were presented to each participant in the same order, grouped according to the acquisition site.

### Evaluation Scores and Performance Measures

Gold Standard annotation of caries was obtained from a panel of 5 dento maxillo-facial radiologists and 1 Professor of Restorative Dentistry, each of whom performed clinical evaluation on a set of images and provided annotation on the location and grade of caries. These individual expert annotations were consolidated resulting in a gold standard set of 1,972 examples of enamel-only proximal caries for algorithm training and evaluation.

The caries annotations entered by each participant were collected remotely and analysed to determine whether they were an Enamel-only True Positive (correct identification of enamel-only proximal caries) or a false positive (an annotation not corresponding to the location of any proximal caries). Annotations corresponding to dentine proximal caries were recorded but excluded from this analysis. Sensitivity of diagnosis is a measure of how well a participant detected enamel-only proximal caries, and calculated as the sum of enamel-only True Positives divided by the sum of enamel-only Gold Standard Caries. The number of False Positives is a measure of how many healthy surfaces the participant incorrectly identified as being carious. It is summed over all images and expressed as the average number of false positives per image.

## Results

In order to compare performance of the control and experimental arms, Table 1 presents a per-participant breakdown of the evaluation scores and performance measures together with the aggregate scores and measures for each arm.

**Table 1.** Per-participant breakdown of the evaluation for each participant and aggregate performance measures for each arm. All but two of the control arm analysed all 24 images, with one failing to analyse 1 image and another only managing to analyse 12. All but one of the experimental arm managed to analyse all images, with the exception failing to analyse 2 of the images. The aggregate measures presented at the bottom of the table are calculated as the sum of the quantitative measures across all participants within each arm, together with the aggregate performance measures.

|  | Images<br>Analysed | Enamel-only Gold<br>Standards | Enamel-only<br>True<br>Positives | False<br>Positives | Sensitivity | False Positive<br>Rate (Per<br>image) |
| --- | --- | --- | --- | --- | --- | --- |
| **Control Arm (without AssistDent)** |  |  |  |  |  |  |
| C1 | 24 | 65 | 28 | 23 | 0.43 | 0.96 |
| C2 | 24 | 65 | 34 | 25 | 0.52 | 1.04 |
| C3 | 24 | 65 | 27 | 15 | 0.42 | 0.63 |
| C4 | 24 | 65 | 9 | 74 | 0.14 | 3.08 |
| C5 | 24 | 65 | 46 | 63 | 0.71 | 2.63 |
| C6 | 12 | 46 | 39 | 103 | 0.85 | 8.58 |
| C7 | 23 | 64 | 23 | 50 | 0.36 | 2.17 |
| C8 | 24 | 65 | 27 | 20 | 0.42 | 0.83 |
| C9 | 24 | 65 | 41 | 69 | 0.63 | 2.88 |
| C10 | 24 | 65 | 30 | 76 | 0.46 | 3.17 |
| C11 | 24 | 65 | 35 | 27 | 0.54 | 1.13 |
| Aggregate | 251 | 695 | 339 | 545 | 0.49 | 2.17 |
| **Experimental Arm (with AssistDent)** |  |  |  |  |  |  |
| E1 | 24 | 65 | 56 | 65 | 0.86 | 2.71 |
| E2 | 24 | 65 | 46 | 50 | 0.71 | 2.08 |
| E3 | 24 | 65 | 52 | 38 | 0.80 | 1.58 |
| E4 | 22 | 63 | 44 | 41 | 0.70 | 1.86 |
| E5 | 24 | 65 | 55 | 58 | 0.85 | 2.42 |
| E6 | 24 | 65 | 55 | 66 | 0.85 | 2.75 |
| E7 | 24 | 65 | 47 | 73 | 0.72 | 3.04 |
| E8 | 24 | 65 | 55 | 122 | 0.85 | 5.08 |
| E9 | 24 | 65 | 58 | 77 | 0.89 | 3.21 |
| E10 | 24 | 65 | 48 | 54 | 0.74 | 2.25 |
| E11 | 24 | 65 | 54 | 59 | 0.83 | 2.46 |
| E12 | 24 | 65 | 53 | 53 | 0.82 | 2.21 |
| Aggregate | 286 | 778 | 623 | 756 | 0.80 | 2.64 |

In order to visualise the distribution of performance measures, Figure 2 illustrates the Sensitivity and False Positive Rate measures of each participant on a scatter plot.

**Figure 2.**
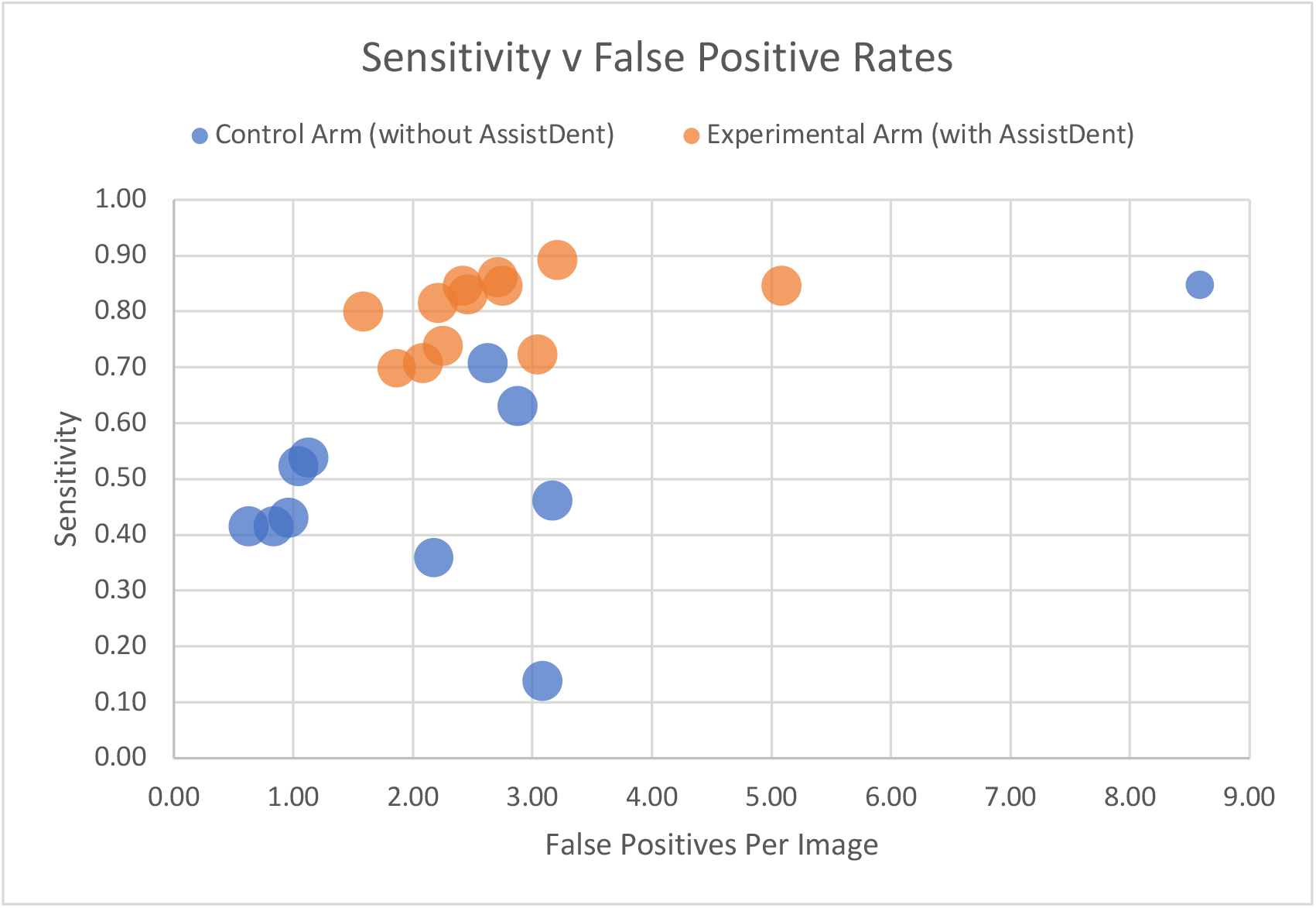
Scatter plot of the Sensitivity versus False Positive Rate for each participant coloured according to arm. The size of each point is representative of the number of images analysed by each participant. The plot illustrates the close grouping of the sensitivity measures for the experimental arm with, on average, improved sensitivity compared to the control arm. The plot also illustrates a similarity in recording False Positives per image by both arms. The outlier to the top right represents a participant within the control arm who failed to analyse half of the images.

Figure 3 presents the mean sensitivity and false positive rates, together with the 95% confidence intervals, for each arm.

**Figure 3.**
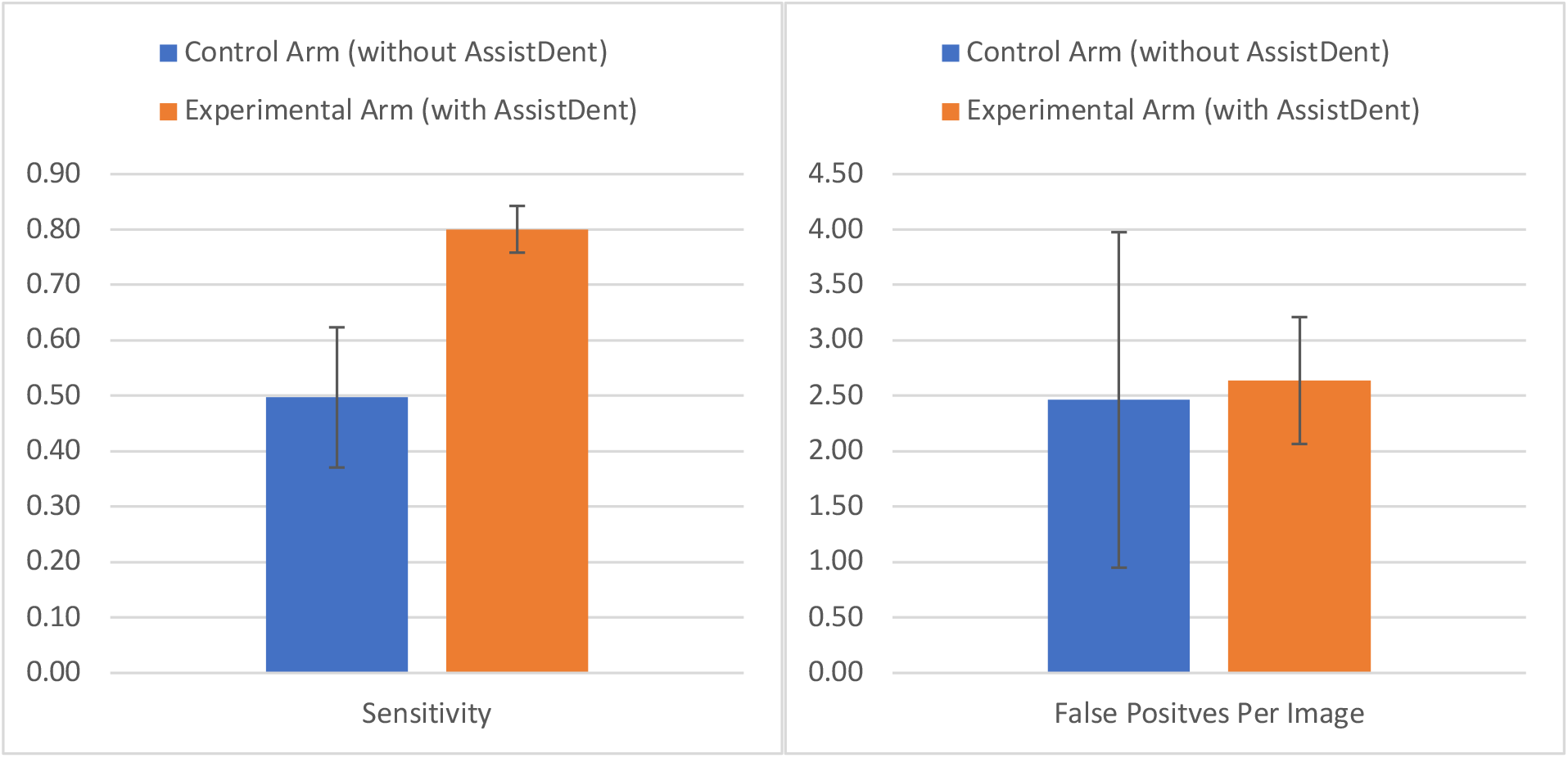
Bar charts showing the mean sensitivities and false positive rates, together with their 95% confidence intervals, for each arm. The improved mean sensitivity of the experimental arm (0.80 with AssistDent) compared to the control arm (0.50 without AssistDent) is clearly visible. The plots show that the 95% confidence interval ranges are narrower in the AssistDent arm compared to the control arm for both measures.

Table 2 presents the statistical analysis for the performance measures within each arm together with the result of a student t-test.

**Table 2.** Statistical analysis of the per-participant performance measures for each arm. The results demonstrate the improved sensitivity of the experimental compared to the control arm with a student t-test p-value well below the usual alpha of 0.01. Results also demonstrate no statistical evidence that the false positive rates are different.

|  | Sensitivity |  | False Positive Per Image |  |
| --- | --- | --- | --- | --- |
|  | Control Arm<br>(Without<br>AssistDent) | Experimental<br>Arm (With<br>AssistDent) | Control Arm<br>(Without<br>AssistDent) | Experimental<br>Arm (With<br>AssistDent) |
| n | 11 | 12 | 11 | 12 |
| Mean | 0.50 | 0.80 | 2.46 | 2.64 |
| 95%CI | 0.13 | 0.04 | 1.51 | 0.57 |
| p-value | 0.00003 |  | 0.81 |  |

In order to highlight outlier images or bias introduced by the order in which the participants analysed the images, Figure 4 presents a per-image breakdown of the average sensitivity for each image for both arms.

**Figure 4.**
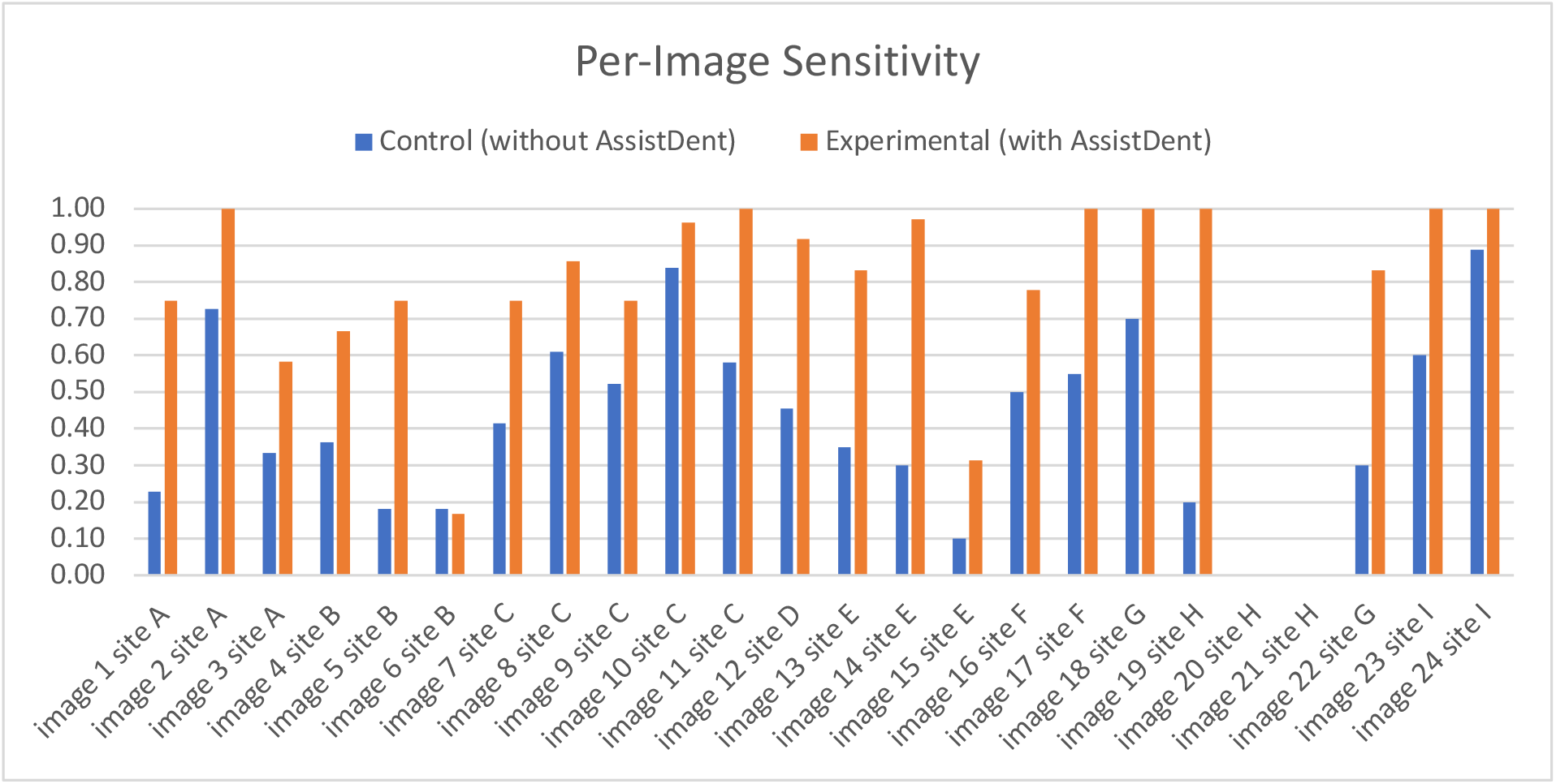
Per-image breakdown of the sensitivity for each arm. Images are numbered (1 to 24) according to the order in which they were presented and grouped by acquisition sites (A to I). The results do not indicate any systematic pattern in the analysis due to the order of presentation of the images to the participants. The graph demonstrates a consistent pattern of improved sensitivity for the experimental arm compared to the control arm for all images except image 6. Note that images 20 and 21 had no enamel-only caries.

## Discussion

In this study we demonstrated that the use of the AI-based software, AssistDent, significantly increased 3^rd^ year students’ ability to detect enamel-only proximal caries in bitewing radiographs. Furthermore, use of the software did not result in significant increased false positive rates. Detecting proximal caries, especially enamel-only lesions, is a difficult clinical task. Digital images can be manipulated for increased clarity, but different dentists may apply different criteria as to whether a carious lesion is present or not. There is often a lack of application of objective criteria, whereas the deterministic AI-based system presented in this paper provides an objective and consistent approach to aid the clinician in formulating a diagnosis. Bader et al. [9] in a classic systematic review found that there was a wide range of proximal caries detected (i.e. 38-94%). For proximal enamel caries, they quoted two studies that had found a mean sensitivity of 41% for visual inspection of radiographs and a mean specificity of 78%. For those patients at high risk of developing caries, highly sensitive methods of caries detection are needed so that preventive regimes can be instituted in an expeditious manner.

The main advantage of using AI-based software is to improve dentists’ performance in the laborious, routine tasks in a cost-effective manner. We have shown that AI can increase the efficiency and safety of detection of enamel caries. Subsequently, when a diagnosis and treatment plan are agreed by the patient and dentist, the process of recording the data in the patient’s notes can be further automated with a resulting saving in time. As well as an aid to student teaching, the AI technology could easily provide a global summary of the patient’s dental health, thereby providing a patient education role. This would allow and empower patients to participate in their dental care.

Our result of 0.80 sensitivity for the detection of enamel-only proximal caries in any tooth of a dental radiograph when users are assisted by AI-based software compares favorably with other published results. Both Jae-Hong Lee [10] and Srivastava [11] reported identical mean sensitivity of 0.81 for their respective fully automated systems, but they measured the detection sensitivity for dentine proximal caries, in addition to enamel caries, which are generally much easier to detect. Valizadeh [12] reported sensitivity of 0.6 for enamel-only caries in their fully automated system.

Our result of 0.50 sensitivity for the detection of enamel-only caries in the control arm of 3^rd^ year dental students, without the use of AssistDent, is comparable with the sensitivity of 0.42 reported for a control arm of qualified dentists reported by Srivastava [11].

AI-based prompting systems are of value in other medical fields in which inspection of radiographs is necessary e.g. cancer screening via mammography images, identifying vertebral fractures in spinal scans, and diagnosing diabetic retinopathy via optical coherence tomography. There is a ready application for AI-based prompting systems in dentistry as dentists frequently use radiographs to diagnose caries, periodontal disease and more. This paper contributes to the emerging field of AI applications in dentistry.

We conclude that the AI-based software, AssistDent, significantly improves the dental students’ ability to detect enamel-only proximal caries and should be considered as a tool to support minimum intervention and preventive dentistry in teaching hospitals and general practice.

## Follow-On Study

This pilot study provided invaluable information in the planning and design of a follow-on study for AssistDent’s use in dental practice. Despite conducting this pilot study under UK lockdown restrictions during the COVID-19 pandemic, we have demonstrated it was possible to comply with the protocol, meet participant recruitment targets and conduct participant sessions remotely. Only minor alterations are proposed for the full study’s protocol and technical support to streamline data acquisition. The pilot study confirmed that it was possible to recruit enough participants from a well-defined and consistent cohort of dental professionals with very similar level of experience. Although some information on how long each participant took to analyse each image was collected during the pilot study, it was identified that the protocol and supporting study technology should be altered to achieve better regulation of this data. It was also recognised that the order in which the images are presented to the participants should be randomised to avoid bias.

It is proposed that a follow-on study will follow a similar design of equally sized control and experimental arms in congruous cohorts of dental students and professionals with different levels of experience to test if use of AssistDent shows similar benefits with more experienced dentists.

## Data Availability

No supplementary data provided

## Notes

### Competing Interest Statement

1. Professor Hugh Devlin is a Professor of Restorative Dentistry in the Division of Dentistry, The University of Manchester and a director of Manchester Imaging Ltd.
2. Manchester Imaging Ltd. received a software licence fee payment from The University of Manchester and the Manchester University NHS Foundation Trust

### Clinical Trial

2019-8534-12770

### Funding Statement

No external funding was received

### Author Declarations

Manchester University Research Ethics Committee (Ref: 2019-8534-12770)

## References

[1] T. Gimenez, C. Piovesan, M. Braga, D. Raggio, C. Deery and D. Ricketts, “Visual inspection for caries detection: a systematic review and meta-analysis,” J. Dent. Res., vol. 94, pp. 895–904, 2015.

[2] F. Schwendicke, M. Tzschoppe and S. Paris, “Radiographic caries detection: a systematic review and meta-analysis,” Journal of dentistry, vol. 43, no. 8, pp. 924–933, 2015.

[3] J. Künisch, G. Schaefer, V. Pitchika, F. Garcia-Godoy and R. Hickel, “Evaluation of detecting proximal caries in posterior teeth via visual inspection, digital bitewing radiography and near-infrared light transillumination.,” American journal of dentistry, vol. 32, no. 2, pp. 74–80, 2019.

[4] M.-A. Geibel, S. Carstens, U. Braisch, A. Rahman, M. Herz and A. Jablonski-Momeni, “Radiographic diagnosis of proximal caries—influence of experience and gender of the dental staff,” Clinical oral investigations, vol. 21, no. 9, pp. 2761–2770, 2017.

[5] E. Pakbaznejad Esmaeili, T. Pakkala, J. Haukka and P. Siukosaari, “Low reproducibility between oral radiologists and general dentists with regards to radiographic diagnosis of caries,” Acta Odontologica Scandinavica, vol. 76, no. 5, pp. 346–350, 2018.

[6] D. A. Young, “New caries detection technologies and modern caries management: merging the strategies.,” General dentistry, vol. 50, no. 4, pp. 320–331, 2002.

[7] S. Tikhonova, F. Girard and M. Fontana, “Cariology education in Canadian dental schools: Where are we? Where do we need to go?,” Journal of dental education, vol. 82, no. 1, pp. 39–46, 2018.

[8] A. C. Leon, L. L. Davis and H. C. Kraemer, “The role and interpretation of pilot studies in clinical research,” Journal of Psychiatric Research, vol. 45, no. 5, pp. 626–629, 5 2011.

[9] J. D. Bader, D. A. Shugars and A. J. Bonito, “Systematic reviews of selected dental caries diagnostic and management methods,” Journal of dental education, vol. 65, no. 10, pp. 960–968, 2001.

[10] J. H. Lee, D. H. Kim, S. N. Jeong and S. H. Choi, “Detection and diagnosis of dental caries using a deep learning-based convolutional neural network algorithm,” Journal of Dentistry, vol. 77, pp. 106–111, 1 10 2018.

[11] M. M. Srivastava, P. Kumar, L. Pradhan and S. Varadarajan, “Detection of tooth caries in bitewing radiographs using deep learning,” arXiv preprint arXiv:1711.07312, 2017.

[12] S. Valizadeh, M. Goodini, S. Ehsani, H. Mohseni, F. Azimi and H. Bakhshandeh, “Designing of a computer software for detection of approximal caries in posterior teeth,” Iranian Journal of Radiology, vol. 12, no. 4, 2015.

